# Single-dose mRNA vaccine effectiveness against SARS-CoV-2, including P.1 and B.1.1.7 variants: a test-negative design in adults 70 years and older in British Columbia, Canada

**DOI:** 10.1101/2021.06.07.21258332

**Authors:** Danuta M Skowronski, Solmaz Setayeshgar, Macy Zou, Natalie Prystajecky, John R Tyson, Eleni Galanis, Monika Naus, David M Patrick, Hind Sbihi, Shiraz El Adam, Bonnie Henry, Linda M N Hoang, Manish Sadarangani, Agatha N Jassem, Mel Krajden

## Abstract

**Introduction:** Randomized-controlled trials of mRNA vaccine protection against SARS-CoV-2 included relatively few elderly participants. We assess singe-dose mRNA vaccine effectiveness (VE) in adults ≥70-years-old in British Columbia (BC), Canada where the second dose was deferred by up to 16 weeks and where a spring 2021 wave uniquely included co-dominant circulation of B.1.1.7 and P.1 variants of concern (VOC).

**Methods:** Analyses included community-dwelling adults ≥70-years-old with specimen collection between April 4 (epidemiological week 14) and May 1 (week 17). Adjusted VE was estimated by test-negative design through provincial laboratory and immunization data linkage. Cases were RT-PCR test-positive for SARS-CoV-2 and controls were test-negative. Vaccine status was defined by receipt of a single-dose ≥21 days before specimen collection, but a range of intervals was assessed. In variant-specific analyses, test-positive cases were restricted to those genetically-characterized as B.1.1.7, P.1 or non-VOC.

**Results:** VE analyses included 16,993 specimens: 1,226 (7.2%) test-positive cases and 15,767 test-negative controls. Of 1,131 (92%) viruses genetically categorized, 509 (45%), 314 (28%) and 276 (24%) were B.1.1.7, P.1 and non-VOC lineages, respectively. VE was negligible at 14% (95% CI 0-26) during the period 0-13 days post-vaccination but increased from 43% (95% CI 30-53) at 14-20 days to 75% (95% CI 63-83) at 35-41 days post-vaccination. VE at ≥21 days was 65% (95% CI 58-71) overall: 72% (95% CI 58-81), 67% (95% CI 57-75) and 61% (95% CI 45-72) for non-VOC, B.1.1.7 and P.1, respectively.

**Conclusions:** A single dose of mRNA vaccine reduced the risk of SARS-CoV-2 in adults ≥70-years-old by about two-thirds, with protection only minimally reduced against B.1.1.7 and P.1 variants. Substantial single-dose protection in older adults reinforces the option to defer the second dose when vaccine supply is scarce and broader first-dose coverage is needed.

## INTRODUCTION

The first mRNA vaccines against COVID-19 (Pfizer-BioNTech; Moderna) were authorized in Canada in December, 2020 [1-3]. In randomized-controlled trials of both products, two doses spaced 3-4 weeks apart were 94-95% efficacious against symptomatic, laboratory-confirmed SARS-CoV-2 infection [2,3]. When RCT data were re-analyzed applying the usual two-week lag for vaccine effect, a single dose of either product was also substantially protective at 92-93% [3,4]. Participants in these trials, however, were generally young and healthy with not more than 5% ≥75-years-old [2,3].

In the context of elevated epidemic activity and scarce vaccine supply, some jurisdictions have extended the interval between first and second SARS-CoV-2 vaccine doses to enable more people to benefit from substantial single-dose protection. In the United Kingdom an interval of up to 12 weeks was recommended on December 30, 2020[5]. In Canada, an even longer interval of up to 16 weeks was recommended beginning March 3, 2021 (epidemiological week 9) [6]. As in most provinces, British Columbia (BC), initially prioritized available mRNA vaccines to long-term care facility (LTCF) residents and frontline healthcare workers. This was associated with dramatic reduction in reported LTCF outbreaks and associated cases [6,7]. However, high vaccine coverage (>90%), including a majority (>60%) who were twice-immunized before week 9 made it difficult to distinguish first-from second-dose and direct from indirect vaccine effects in that relatively closed setting.

Community vaccination in BC subsequently followed an age-based strategy that first prioritized older adults ≥90, 80-89 and 70-79 years beginning around week 10. Although viral vector vaccines are also authorized in Canada [1], they were not prominently used in these age groups. In the spring 2021, BC experienced its most substantial pandemic wave to date, including a majority of viruses that were characterized as variants of concern (VOC), and uniquely including co-dominant circulation of P.1 and B.1.1.7 [9]. A publicly-funded, mostly symptom-based approach for SARS-CoV-2 diagnostic testing is broadly accessible in BC. In that context, we applied a test-negative design (TND) to estimate the vaccine effectiveness (VE) of a single dose of mRNA vaccine against SARS-CoV-2, including variant-specific estimates, among community-dwelling adults ≥70-years-old in BC.

## METHODS

### Source population, analysis period and study design

There are about 673,000 adults ≥70-years-old in BC (13% of the total 5.1 million population) including ∼437,000 (65%) 70-79 years, 188,000 (28%) 80-89 years and 48,000 (7%) ≥90 years old with slightly more than half who are women (54%) [8].

The spring 2021 wave peaked in BC in week 14 and gradually subsided with province-wide restrictions; however, weekly case reports continued to exceed the peak week of prior waves until week 17 [7]. The analysis period of the current study spanned weeks 14 to 17 (April 4-May 1), taking into account vaccine roll-out and several-week delay for vaccine effect as well as community SARS-CoV-2 activity that remained elevated during this period.

VE was assessed by TND with multivariable logistic regression used to estimate the adjusted odds ratio (OR_adj_) for vaccination among test-positive cases versus test-negative controls. VE and 95% confidence intervals (CI) were derived as (1-OR_adj_) x 100%. The following covariates were included in adjusted models: age group, sex, epidemiological week and health authority (HA) of residence, or if the latter were not available then the HA of the clinician associated with the test.

### Data sources

Specimens collected between weeks 14-17 and tested by RT-PCR for SARS-CoV-2 were eligible. Test-positive cases and test-negative controls were sampled from within the Public Health Laboratory Operations Viewer and Reporter (PLOVER) database. PLOVER was established by the BC Centre for Disease Control (BCCDC) Public Health Laboratory (PHL) to capture, in real time, all specimens tested for SARS-CoV-2 along with client, specimen collection and testing details; however, symptoms and onset date are not consistently captured in PLOVER. Vaccination information was obtained from the provincial immunization registry (PIR), a centralized database that captures, also in real-time, all SARS-CoV-2 vaccinations in BC, along with client and vaccination details. Individual-level linkage between PLOVER and PIR datasets was achieved through unique personal identifiers.

### Case and control selection

Individuals could contribute a single test-positive specimen. In variant-specific analyses, test-positive cases were restricted to those in whom a VOC was detected, classified as P.1, B.1.1.7 or non-VOC as defined in **Supplementary Material S1**. Three approaches were used for test-negative control selection. In the first specimen-based approach, all negative specimens from a single individual could contribute; however, specimens collected on the same day were counted only once or excluded if discordant. In the second individual-based approach, only the single latest negative specimen per individual could contribute. In an alternative individual-based approach, only one randomly-selected negative specimen per individual could contribute. We further explored with and without exclusion of negative specimens collected within three weeks before a positive specimen.

### Vaccine status definition

Clients with record of a single dose of mRNA vaccine on or before the date of specimen collection were considered vaccinated; those without such record were considered unvaccinated. Because our VE analyses are timed on specimen collection rather than onset date we incorporate additional lag beyond the usual two week grace period for vaccine effect. Among community-dwelling adults ≥70-years-old with both dates available in PLOVER, the mean and median interval between onset and specimen collection date was 4 and 3 days, respectively, with interquartile range of 1-5 days. We base primary VE analyses on vaccine receipt at least three weeks before specimen collection date (≥21 days) but assess intervals of 0-13, 14-20, 21-27, 28-41 and ≥42 days.

### Inclusion/exclusion criteria

Specimens missing information for age, sex, HA, specimen collection date, vaccine date or type were excluded as were those with missing or inconclusive RT-PCR results. Cases with collection date before the start of the analysis period were excluded, identified through further linkage with the notifiable disease list of confirmed COVID-19 cases reported by the HAs and maintained by the BCCDC. Specimens that were tested outside of public funding were excluded because of systematically lower likelihood of test-positivity [7]. Clients who received more than one vaccine dose were excluded as were those who received a viral vector vaccine [1]. Finally, any specimens identified within PLOVER and/or the PIR or notifiable disease list from LTCF, assisted-living or independent-living facilities were excluded.

### Ethics statement

Data linkages and analyses were conducted under a surveillance mandate, authorized by the Provincial Health Officer under the Public Health Act, and exempt from research ethics board review.

## RESULTS

### Participant profiles

In total, 16,993 SARS-CoV-2 specimens contributed to VE analyses, including 1,226 (7.2%) test-positive cases and 15,767 test-negative controls (**Figure S2**). Viruses from 1,131/1,226 (92%) cases were genetically categorized with respect to VOC status, of which 509 (45%) and 314 (28%) were B.1.1.7 and P.1, respectively (**Tables S1, S6; and S7**). An additional 4 (<1%) viruses belonged to the B.1.351 lineage and another 12 (1%) could not be differentiated as P.1 or B.1.351 while 16 viruses (1%) were B.1.617.1/2; these were excluded from variant-specific VE analyses (**Table S1**). Of the remainder, 276 (24%) were designated non-VOC. The distributions of VOC and non-VOC by participant sub-group were similar (**Figure S1**).

Decrease in test-positivity and case tallies by successive week of the analysis period mirrored provincial surveillance patterns (**Figure 1; Table 1**) [7]. The distributions of test-negative controls by age, sex and HA were generally representative of the BC source population (**Table 1**) [7,8].

**Table 1.**
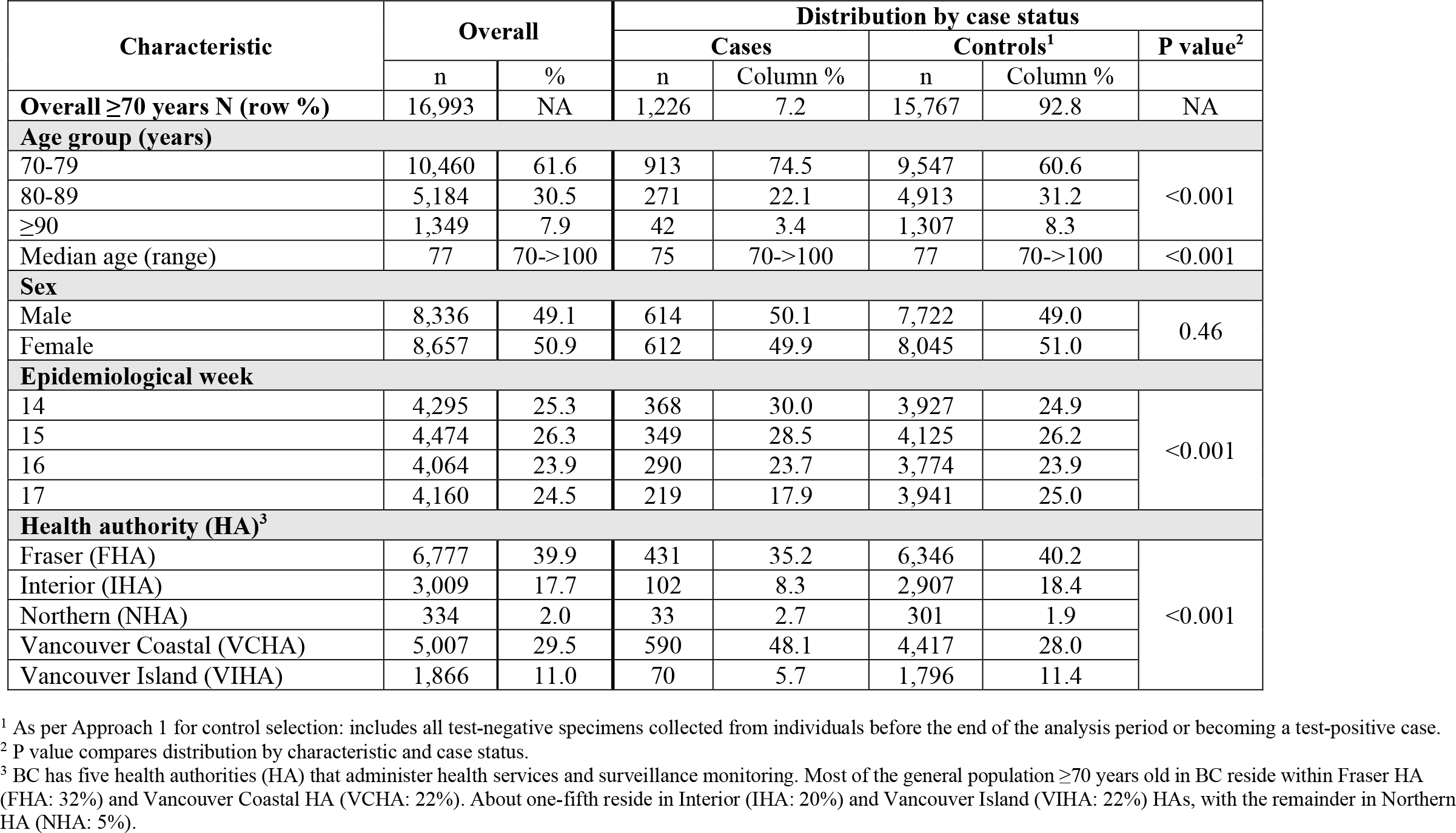
Participant characteristics by case and control status, adults ≥70 years of age, British Columbia (BC), Canada, weeks 14-17

| Characteristic | Overall |  | Distribution by case status |  |  |  |  |
| --- | --- | --- | --- | --- | --- | --- | --- |
|  |  |  | Cases |  | Controls<sup>1</sup> |  | P value<sup>2</sup> |
|  | n | % | n | Column % | n | Column % |  |
| Overall ≥70 years N (row %) | 16,993 | NA | 1,226 | 7.2 | 15,767 | 92.8 | NA |
| Age group (years) |  |  |  |  |  |  |  |
| 70-79 | 10,460 | 61.6 | 913 | 74.5 | 9,547 | 60.6 | <0.001 |
| 80-89 | 5,184 | 30.5 | 271 | 22.1 | 4,913 | 31.2 |  |
| ≥90 | 1,349 | 7.9 | 42 | 3.4 | 1,307 | 8.3 |  |
| Median age (range) | 77 | 70->100 | 75 | 70->100 | 77 | 70->100 | <0.001 |
| Sex |  |  |  |  |  |  |  |
| Male | 8,336 | 49.1 | 614 | 50.1 | 7,722 | 49.0 | 0.46 |
| Female | 8,657 | 50.9 | 612 | 49.9 | 8,045 | 51.0 |  |
| Epidemiological week |  |  |  |  |  |  |  |
| 14 | 4,295 | 25.3 | 368 | 30.0 | 3,927 | 24.9 | <0.001 |
| 15 | 4,474 | 26.3 | 349 | 28.5 | 4,125 | 26.2 |  |
| 16 | 4,064 | 23.9 | 290 | 23.7 | 3,774 | 23.9 |  |
| 17 | 4,160 | 24.5 | 219 | 17.9 | 3,941 | 25.0 |  |
| Health authority (HA)<sup>3</sup> |  |  |  |  |  |  |  |
| Fraser (FHA) | 6,777 | 39.9 | 431 | 35.2 | 6,346 | 40.2 | <0.001 |
| Interior (IHA) | 3,009 | 17.7 | 102 | 8.3 | 2,907 | 18.4 |  |
| Northern (NHA) | 334 | 2.0 | 33 | 2.7 | 301 | 1.9 |  |
| Vancouver Coastal (VCHA) | 5,007 | 29.5 | 590 | 48.1 | 4,417 | 28.0 |  |
| Vancouver Island (VIHA) | 1,866 | 11.0 | 70 | 5.7 | 1,796 | 11.4 |  |

<sup>1</sup> As per Approach 1 for control selection: includes all test-negative specimens collected from individuals before the end of the analysis period or becoming a test-positive case.<sup>2</sup> P value compares distribution by characteristic and case status.<sup>3</sup> BC has five health authorities (HA) that administer health services and surveillance monitoring. Most of the general population $\geq 70$ years old in BC reside within Fraser HA (FHA: 32%) and Vancouver Coastal HA (VCHA: 22%). About one-fifth reside in Interior (IHA: 20%) and Vancouver Island (VIHA: 22%) HAs, with the remainder in Northern HA (NHA: 5%).

**Figure 1.**
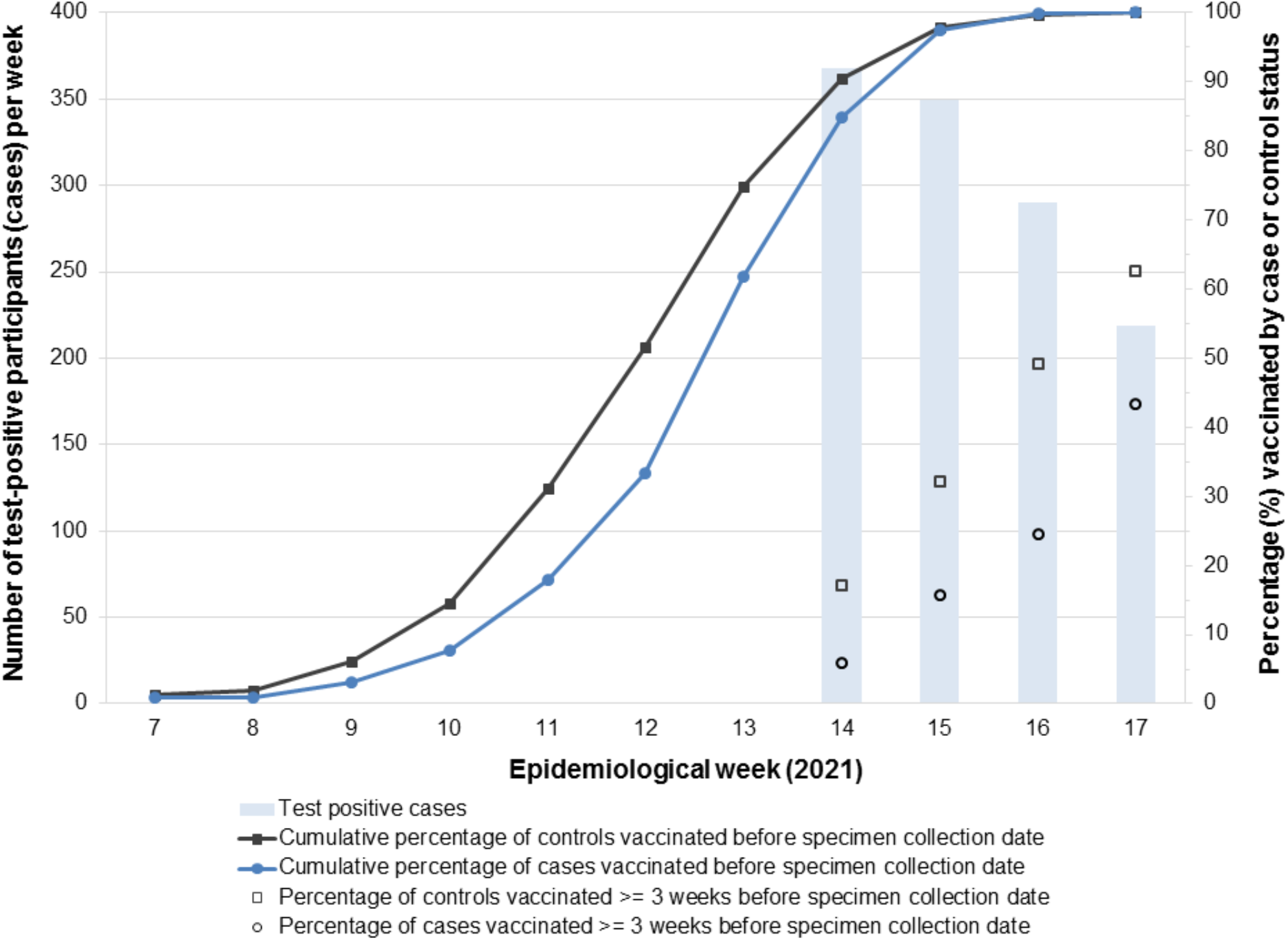
Percentage vaccinated among SARS-CoV-2 test-positive cases and test-negative controls and case tallies by epidemiological week, participating adults ≥70 years of age, British Columbia, Canada, weeks 14-17

Among vaccinated cases and controls, 85% and 90%, respectively, had received their first dose by week 14 (**Figure 1**). Among test-negative controls, vaccine coverage was comparable to the provincial average for community-dwelling adults ≥70 years overall (74% vs. 75%), and by week 14 (60% vs. 64%), 15 (72% vs. 75%), 16 (82% vs. 80%) and 17 (84% vs. 82%) (**Table 2**). Of specimens from vaccinated cases and controls, >90% were collected <42 days since vaccination, limiting VE interpretation after that period. Most (85%) vaccinated individuals had received the Pfizer-BioNTech product.

**Table 2.** Participant characteristics by vaccine status, adults ≥70 years of age, British Columbia (BC), Canada, weeks 14-17

| Characteristic | Number of participants |  |  | Number and percent vaccinated by characteristic and case status <sup>1</sup> |  |  |  |  |  |  |
| --- | --- | --- | --- | --- | --- | --- | --- | --- | --- | --- |
|  | Overall | Cases | Controls<sup>2</sup> | Overall (n, %) |  | P value<sup>3</sup> | Cases (n, %) |  | Controls<sup>2</sup> (n, %) |  |
| Overall ≥70 years | 16,993 | 1,226 | 15,767 | 12,451 | 73.3 | NA | 751 | 61.3 | 11,700 | 74.2 |
| Age group (years) (row percentages displayed) |  |  |  |  |  |  |  |  |  |  |
| 70-79 | 10,460 | 913 | 9,547 | 7,073 | 67.6 | <0.001 | 529 | 57.9 | 6,544 | 68.5 |
| 80-89 | 5,184 | 271 | 4,913 | 4,279 | 82.5 |  | 191 | 70.5 | 4,088 | 83.2 |
| ≥90 | 1,349 | 42 | 1,307 | 1,099 | 81.5 |  | 31 | 73.8 | 1,068 | 81.7 |
| Median age | 77 | 75 | 77 | 78 | 70->100 | >0.05 | 76 | 70-99 | 78 | 70->100 |
| Sex |  |  |  |  |  |  |  |  |  |  |
| Male | 8,336 | 614 | 7,722 | 6,095 | 73.1 | 0.02 | 386 | 62.9 | 5,709 | 73.9 |
| Female | 8,657 | 612 | 8,045 | 6,356 | 73.4 |  | 365 | 59.6 | 5,991 | 74.5 |
| Epidemiological week of specimen collection (row percentages displayed) |  |  |  |  |  |  |  |  |  |  |
| 14 | 4,295 | 368 | 3,927 | 2,532 | 59.0 | <0.001 | 180 | 48.9 | 2,352 | 59.9 |
| 15 | 4,474 | 349 | 4,125 | 3,172 | 70.9 |  | 210 | 60.2 | 2,962 | 71.8 |
| 16 | 4,064 | 290 | 3,774 | 3,303 | 81.3 |  | 208 | 71.7 | 3,095 | 82.0 |
| 17 | 4,160 | 219 | 3,941 | 3,444 | 82.8 |  | 153 | 69.9 | 3,291 | 83.5 |
| Health authority (HA) (row percentages displayed) |  |  |  |  |  |  |  |  |  |  |
| Fraser (FHA) | 6,777 | 431 | 6,346 | 5,119 | 75.5 | <0.001 | 242 | 56.1 | 4,877 | 76.9 |
| Interior (IHA) | 3,009 | 102 | 2,907 | 2,072 | 68.9 |  | 56 | 54.9 | 2,016 | 69.3 |
| Northern (NHA) | 334 | 33 | 301 | 222 | 66.5 |  | 20 | 60.6 | 202 | 67.1 |
| Vancouver Coastal (VCHA) | 5,007 | 590 | 4,417 | 3,791 | 75.7 |  | 397 | 67.3 | 3,394 | 76.8 |
| Vancouver Island (VIHA) | 1,866 | 70 | 1,796 | 1,247 | 66.8 |  | 36 | 51.4 | 1,211 | 67.4 |
| Vaccine product (column percentages displayed) |  |  |  |  |  |  |  |  |  |  |
| Pfizer BioNTech | NA | NA | NA | 10,569 | 84.9 | NA | 646 | 86.0 | 9,923 | 84.8 |
| Moderna |  |  |  | 1,882 | 15.1 |  | 105 | 14.0 | 1,777 | 15.2 |
| Days since vaccination (DSV)<sup>4</sup> (column percentages displayed) |  |  |  |  |  |  |  |  |  |  |
| 0-13 | NA | NA | NA | 3,432 | 27.6 | NA | 345 | 45.9 | 3,087 | 26.4 |
| 14-20 |  |  |  | 2,464 | 19.8 |  | 163 | 21.7 | 2,301 | 19.7 |
| 21-27 |  |  |  | 2,302 | 18.5 |  | 110 | 14.6 | 2,192 | 18.7 |
| 28-34 |  |  |  | 1851 | 14.9 |  | 61 | 8.1 | 1790 | 15.3 |
| 35-41 |  |  |  | 1210 | 9.7 |  | 30 | 4.0 | 1180 | 10.1 |
| 42-99 |  |  |  | 1192 | 9.6 |  | 42 | 5.6 | 915 | 9.8 |
| Median DSV (range) | NA | NA | NA | 21 | 0-99 | NA | 14 | 0-82 | 22 | 0-99 |

<sup>1</sup> Single-dose recipients only without regard to interval between vaccination and specimen collection. Includes mRNA vaccine receipt only; viral vector vaccine recipients excluded.<sup>2</sup> As per Approach 1 for control selection: includes all test-negative specimens.<sup>3</sup> P value compares percentage vaccinated by characteristic.<sup>4</sup> Interval between first dose of vaccine and specimen collection date

### VE estimates

VE estimates did not vary by the approach used to select test-negative controls and we therefore present VE based on all-specimen inclusion (approach 1) (**Table S2**). VE findings are illustrated in **Figure 2** with details in **Tables S2**-**S7**.

**Figure 2.**
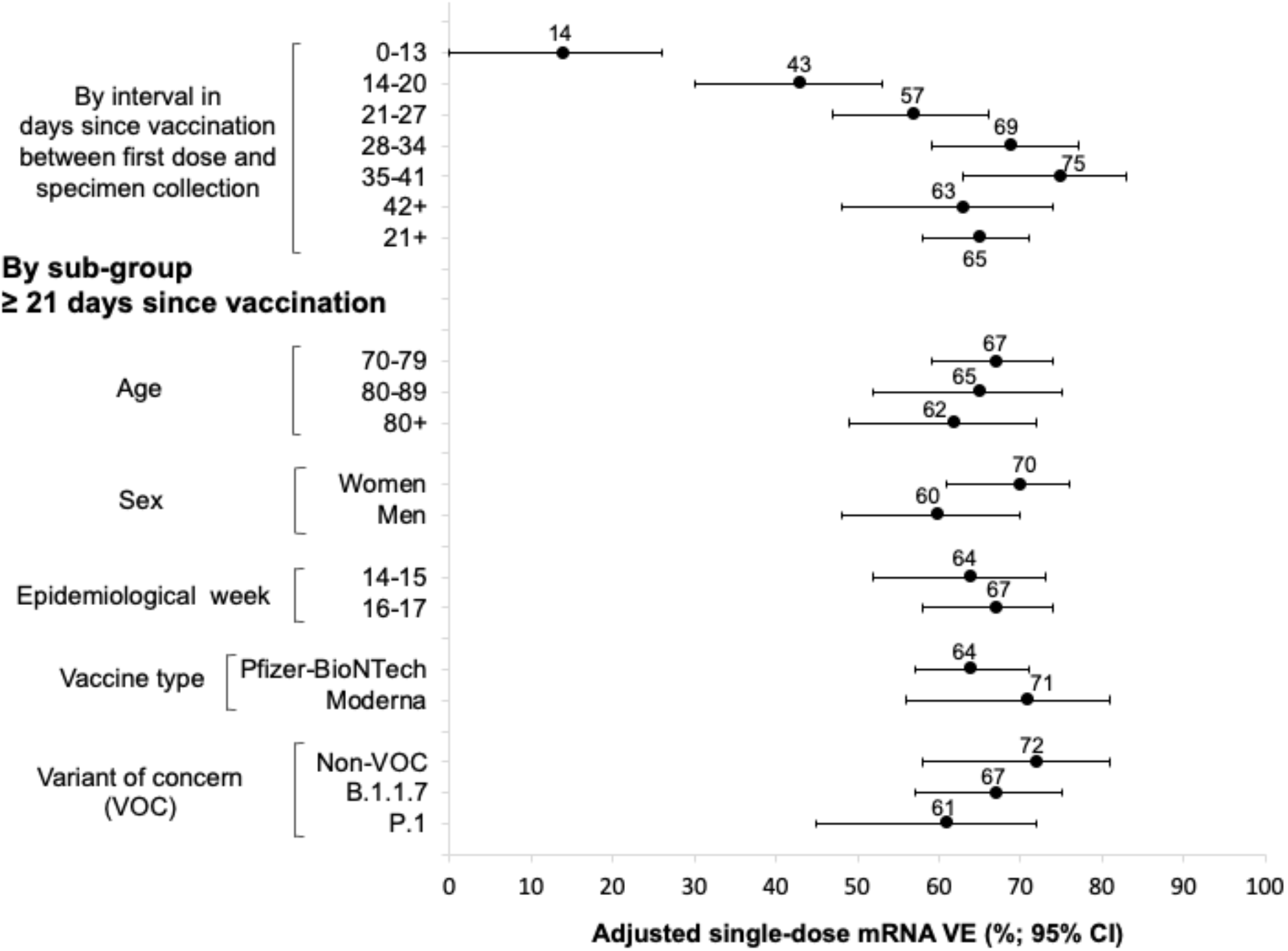
Adjusted vaccine effectiveness estimates by interval in days since vaccination and restricted by sub-group, adults ≥70 years of age, British Columbia, Canada, weeks 14-17 VE = vaccine effectiveness; CI = confidence interval All vaccine effectiveness estimates are adjusted for age group (70-79, 80-89, 90+ years); sex (men, women); epidemiological week (14, 15, 16, or 17); and health authority (HA) (Fraser HA, Interior HA, Northern HA, Vancouver Coastal HA, Vancouver Island HA). See **Supplementary Tables S2-S8** for details.

VE was negligible at 14% (95% CI 0-26) during the period 0-13 days post-vaccination but increased by one week interval thereafter from 43% (95% CI 30-53) at 14-20 days to 75% (95% CI 63-83) at 35-41 days post-vaccination (**Figure 2)**. VE is also displayed for ≥42 days but warrants cautious interpretation given a minority of vaccinated participants belonged within that extended interval (**Table 1**). Summary VE at ≥21 days was 65% (95% CI 58-71) and was similar (within 10% absolute) in participant sub-group analyses, differing by 10% in women (70%; 95% CI 61-76) vs. men (60%; 95% CI 48-70) (**Figure 2; Tables S2-S5**).

At ≥21 days since vaccination, a single dose of mRNA vaccine was also significantly protective in variant-specific analyses, with VE of 72% (95% CI 58-81), 67% (95% CI 57-75) and 61% (95% CI 45-72) for non-VOC, B.1.1.7 and P.1, respectively.

## DISCUSSION

We report substantial protection provided by a single dose of mRNA vaccine against SARS-CoV-2 infection in adults ≥70-years-old. VE increased when longer intervals were used to define vaccine status, becoming statistically significant at 40% after a two-week lag, 60% after three-week, 70% after four-week and 75% after 5-week interval between vaccination and specimen collection. While delayed immunological response in the elderly may be hypothesized to explain this prolonged timeline to protection [10], a methodological explanation also exists, namely misclassification of cases as vaccine-preventable at too-short intervals when based on specimen collection rather than onset date. We underscore the need for studies to extend the interval used to define vaccine status when outcomes are timed on events such as specimen collection or testing that occur later or with more variability than the typical two-week interval from vaccination to onset date used in clinical trials. Our primary VE estimate of 65% based on RT-PCR detection of infection at ≥3 weeks between vaccination and specimen collection may also be an under-estimate. Our findings suggest, however, that a single dose of mRNA vaccine prevented about two out of three SARS-CoV-2 infections in older adults. Such protection is particularly meaningful considering that it was provided during a period of peak pandemic risk, when VOCs were predominantly contributing to the epidemic in BC.

Our VE estimates were robust in sensitivity and subgroup analyses, varying only by about 10% (absolute) based on sex (10% lower in men) and VOC (11% lower for P.1 versus non-VOC). With overlapping confidence intervals, these comparisons are not definitive but signal the need for further evaluation, notably in younger adults among whom sex differences may be more biologically-mediated [11], and VOC circulation more prominent [9]. In BC, where P.1 and have uniquely co-dominated during a substantial spring wave [9], the finding of their comparable VE in older adults is important. This observation aligns well with immunogenicity findings elsewhere reporting comparable reductions in infection- and vaccine-induced neutralizing antibody for P.1 and B.1.1.7 [12]. Whereas more severe reductions in immunity or effectiveness have been reported for other VOC such as B.1.351 or B.1.617 [12-14], we had too-few detections for their separate VE analysis here. Despite some shared substitutions such as E484K between P.1 and B.1.351, they may not be equal in their potential for vaccine escape. To better correlate molecular markers with immunological and epidemiological measures of vaccine protection, and to inform the need for vaccine update, VE analyses should be stratified as finely as possible by genetic sub-cluster.

Our findings may be compared to other similar studies in older adults although underlying differences (e.g. methods, populations, vaccine status and outcome definitions, mix of circulating viruses) need to be taken into account. Using the TND to assess VE among adults ≥70 years in England (but including care-home residents), Bernal et al. reported single-dose mRNA VE against symptomatic SARS-CoV-2 infection reaching 61% (95% CI 51-69) by 28-34 days [15], similar to our estimate of 69% (95% CI 59-77) by the same interval. In a matched case-control study of adults 80-83-years-old in England (excluding care-home residents), Mason et al report (in pre-print) mRNA VE against SARS-CoV-2 infection of 55% (95% CI 41-67) by 21-27 days and 70% (95% CI 55-80) by 35-41 days [16], also similar to our estimates among adults 80-89-years-old of 54% (95% CI 32-70) and 75% (95% CI 55-86), respectively, at those intervals. In a recent pre-print also from Canada, Chung et al use the TND to assess mRNA VE against symptomatic infection for the population of Ontario with primary analysis based on an interval of ≥14 days between vaccination and specimen collection [17]. In sub-analysis of adults ≥70 years (excluding care-home residents), authors report VE of 40% (95% CI 29-49) which is lower than our estimate of 58% (95% CI 50-64) at ≥14 days (not displayed) or our primary analysis of 65% (95% CI 58-71) at ≥21 days. Using an interval of 21-27 days and 28-34 days, however, Chung et al report VE of 40% (95% CI 21-54) and 64% (95% CI 46-76), respectively, the latter being more compatible with other estimates above. Of note, the Ontario analysis spanned mid-December to mid-April but as in BC most of their participants, including those ≥70-years-old, would not have been vaccine-eligible until the tail end of their analysis period, notwithstanding earlier case and control contribution.

Given both time-varying vaccine coverage and disease risk, adjustment for confounding by calendar-time is critical in observational study designs. To address that concern, we restricted our analysis to a narrow window (weeks 14-17) when vaccine coverage and community risk were both high and relatively stable, further adjusting by epidemiological week to address variation. We also explored several approaches for selecting test-negative controls with similar results, also likely reflecting the narrow analysis period we chose. The main limitation of our analysis, as elsewhere, is our reliance on general laboratory submissions and clinical or surveillance data that were originally collected for a different purpose and are subject to missing information and misclassification, as well as selection bias. Although foremost symptom-based, the clinical testing indications for COVID-19 are broad, discretionary and variable. To attempt standardization of the likelihood of test-positivity among sampled specimens we excluded those identified as having been collected from congregate settings or for non-clinical screening purposes. Such exclusions, however, may have been incomplete or introduced other unintended biases. We were limited in the covariates we could include in our model and cannot rule out residual bias and confounding. As a form of validity check, we assessed VE during the 0-13-day period when little or no vaccine effect is anticipated, confirming negligible VE as expected. For similar reasons, we compared vaccine coverage and other characteristics of our test-negative controls to that of the general source population ≥70-years-old in BC, and this was reassuringly concordant. Our findings also align well with other observational studies in older adults each of which are, however, subject to similar issues. Because the PLOVER database from which we sampled does not reliably capture symptoms or onset dates, we assessed VE against any infection without symptom or severity specification. VE estimates against more severe outcomes are anticipated to be higher than we report for infection per se [15-17]. Finally, we were limited in our ability to assess VE over the long-term or to compare to younger age groups prioritized later for vaccination, but those analyses are underway.

In conclusion, a single dose of mRNA vaccine reduced the risk of SARS-CoV-2 infection by about two-thirds in community-dwelling adults ≥70-years-old. Such protection is particularly important because it was observed during a period of peak pandemic risk when VOC, predominantly the B.1.1.7 and P.1 lineages, comprised at least 70% of characterized viruses. Substantial single-dose protection in older adults reinforces the option to defer second doses when vaccine supply is scarce and broader first-dose coverage is needed.

## Data Availability

To the extent they comply with relevant privacy legislation, data sharing will be considered upon request.

## Acknowledgments

Authors thank the following from the BC Centre for Disease Control: Chris Fjell for his leadership and management of the Public Health Laboratory Operations Viewer and Reporter (PLOVER); Yayuk Joffres for quality control of laboratory data; Yin Chang for laboratory data management; May Ahmed for her surveillance-related insights; Shinhye Kim for research coordination support and Samantha Kaweski for laboratory coordination support. We thank the Provincial Public Health Information Systems (PPHIS) team for contributions related to the provincial immunization registry (PIR). We thank the many frontline, regional and provincial practitioners, including clinical, laboratory and public health providers, epidemiologists, Medical Health Officers, laboratory staff, vaccinators and others who provided the epidemiological, virological and genetic characterization data underpinning these analyses.

## Funding

Funding was provided in part by the Michael Smith Foundation for Health Research.

## Potential conflicts of interest

DMS is Principal Investigator on grants from the Michael Smith Foundation for Health Research in support of this work. MK received grants/contracts paid to his institution from Roche, Hologic and Siemens, unrelated to this work. MS is supported via salary awards from the BC Children’s Hospital Foundation, the Canadian Child Health Clinician Scientist Program and the Michael Smith Foundation for Health Research. MS has been an investigator on projects funded by GlaxoSmithKline, Merck, Pfizer, Sanofi-Pasteur, Seqirus, Symvivo and VBI Vaccines. All funds have been paid to his institute, and he has not received any personal payments. Other authors have no conflicts of interest to disclose.

## Notes

### Clinical Trial

Not applicable

### Author Declarations

Data linkages and analyses were conducted under a surveillance mandate, authorized by the Provincial Health Officer under the Public Health Act, and exempt from research ethics review as waived by the University of British Columbia Clinical Research Ethics Board.

## REFERENCES

1. National Advisory Committee on Immunization (NACI). Recommendations on the use of COVID-19 vaccines. Ottawa: NACI. [Accessed 3 June 2021]. Available from: https://www.canada.ca/en/public-health/services/immunization/national-advisory-committee-on-immunization-naci/recommendations-use-covid-19-vaccines.html#a12

2. Polack FP, Thomas SJ, Kitchin N, Absalon J, Gurtman A, Lockhart S, et al. Safety and efficacy of the BNT162b2 mRNA Covid-19 vaccine. N Eng J Med. 2020; 383:2603–2615. https://doi.org/10.1056/NEJMoa203457 PMID: 33301246

3. Baden LR, El Sahly HM, Essink B, Kotloff K, Frey S, Novak R, et al. Efficacy and safety of the mRNA-1273 SARS-CoV-2 vaccine. N Eng J Med. 2020; 384(5):403–416. https://doi.org/10.1056/JENJoa2035389 PMID: 33378609

4. Skowronski DM, De Serres G. Safety and efficacy of the BNT162b2 mRNA Covid-19 vaccine. N Eng J Med. 2021;384(11):10.1056/NEJMc2036242#sa1. https://doi.org/10.1056/NEJMc2036242 PMID: 33596348

5. Joint Committee on Vaccination and Immunisation (JCVI). Joint Committee on Vaccination and Immunisation: advice on priority groups for COVID-19 vaccination. JCVI; 20 Dec 2020. [Accessed 15 May 2021]. Available from: https://assets.publishing.service.gov.uk/government/uploads/system/uploads/attachment_data/file/950113/jcvi-advice-on-priority-groups-for-covid-19-vaccination-30-dec-2020-revised.pdf

6. National Advisory Committee on Immunization (NACI). NACI rapid response: extended dose intervals for COVID-19 vaccines to optimize early vaccine rollout and population protection in the context of limited vaccine supply. Canada. Ottawa: NACI. [Accessed 15 May 2021]. Available from: https://www.canada.ca/en/public-health/services/immunization/national-advisory-committee-on-immunization-naci/extended-dose-intervals-covid-19-vaccines-early-rollout-population-protection.html

7. British Columbia Centre for Disease Control (BCCDC). British Columbia COVID-19 situation report. Vancouver: BCCDC. [Accessed 15 May 2021]. Available from: http://www.bccdc.ca/health-info/diseases-conditions/covid-19/data

8. Statistics Canada. Table 17-10-0005-01. Population estimates on July 1^st^, by age and sex. DOI: https://doi.org/10.25318/1710000501-eng. [Accessed 17 May 2021]. Available from: https://www150.statcan.gc.ca/t1/tbl1/en/tv.action?pid=1710000501

9. British Columbia Centre for Disease Control (BCCDC). COVID-19 VoC report. [Accessed 2 June 2021]. Available from: http://www.bccdc.ca/health-info/diseases-conditions/covid-19/data#variants

10. Boraschi D, Aguado MT, Dutel C, et al. The gracefully aging immune system. Sci. Transl. Med 2013;5:185ps8. Doi: 10.1126/scitranslmed.3005624

11. Klein SL and Pekosz A. Sex-based biology and the rational design of influenza vaccination strategies. J Infect Dis 2014;209(S3):S114–9.

12. Dejnirattisai W, Zhou D, Supasa P, et al. Antibody evasion by the P.1 strain of SARS-CoV-2. Cell 2021;184:2939–54.e9.

13. Vidal SJ, Collier AY, Yu J, et al. Correlates of neutralization against SARS-CoV-2 variants of concern by early pandemic sera. J Virol 2021 Apr 23;JVI.00404-21. Doi: 10.1128/JVI.00404-21. Online ahead of print.

14. Bernal JL, Andrews N, Gower C, et al. Effectiveness of COVID-19 vaccines against the B.1.617.2 variant. medRxiv. 2021. Preprint doi: https://doi.org/10.1101/2021.05.22.21257658

15. Bernal JL, Andrews N, Gower C, et al. Effectiveness of the Pfizer-BioNTech and Oxford-AstraZeneca vaccines on covid-19 related symptoms, hospital admissions, and mortality in older adults in England: test negative case-control study. BMJ 2021;373:n1088

16. Mason T, Whitston M, Hodgson J, et al. Effects of BNT162b2 mRNA vaccine on COVID-19 infection and hospitalization among older people: matched case control study for England. medRxiv. 2021. Preprint doi: https://doi.org/10.1101/2021.04.19.21255461

17. Chung H, He S, Nasreen S, et al. Effectiveness of BnT162b2 and mRNA-1273 COVID-19 vaccines against symptomatic SARS-CoV-2 infection and severe COVID-19 outcomes in Ontario, Canada. medRXiv. 2021. Preprint doi: https://doi.org/10.1101/2021.05.24.21257744

