## Supplementary Material for "Single-dose mRNA vaccine effectiveness against SARS-CoV-2, including P.1 and B.1.1.7 variants: a test-negative design in adults 70 years and older in British Columbia, Canada"

**Supplementary Material S1** Genetic characterization of SARS-CoV-2 viruses

To understand the mix of SARS-CoV-2 viruses underpinning vaccine effectiveness (VE) findings and to support variant-specific VE estimates, we attempted to genetically characterize all viruses collected from the 1,226 cases ≥70 years old who contributed to VE analyses.

Viruses were genetically grouped based upon: (a) whole genome sequencing (WGS) and/or (b) real-time reverse transcription polymerase chain reaction (RT-PCR) screening assays targeting signature mutations of particular variants of concern (VOC)[1,2] Where WGS and screening RT-PCR assays were discordant, WGS results were accepted. Where uncertainty remained, specimens were excluded.

Whole genome sequencing
WGS was performed at the British Columbia Centre for Disease Control Public Health Laboratory (BCCDC PHL). In brief, SARS-CoV-2 RNA was extracted using the Applied BioSystems MagMax™ Express 96 Nucleic Acid Extractor and the MagMax Viral/Pathogen Nucleic Acid Isolation Kit (Thermo Fisher). Viral RNA was reverse transcribed into cDNA using Thermo SuperScript IV. A 1200bp tiled amplicon scheme[3] was used to amplify the entire SARS-CoV-2 genome[4]. DNA libraries were prepared using DNA Prep Library Preparation Kit (Illumina) and sequenced on a MiSeq or NextSeq 2000 instrument (Illumina, San Diego). SARS-CoV-2 whole genome consensus sequences were generated using a modified Nextflow pipeline for running the ARTIC network's[5] field bioinformatics tools[6]. Lineages were assigned using Pangolin (version 2.4.2, pangoLEARN 2021-05-19)[7] and sequencing quality control metrics and mutational profiles used nCoV-tools from the Simpson Lab[8].

Screening RT-PCR
Four VOC screening RT-PCR assays were used at the BCCDC PHL and various hospital-based laboratories across the province including (1) a laboratory-developed N501Y single nucleotide polymorphism (SNP) PCR at the Victoria General Hospital located within Vancouver Island Health Authority[9]; (2) a laboratory-developed N501Y and E484K dual SNP PCR at the BCCDC PHL; (3) a laboratory developed sequential deletion and SNP PCR including N501Y and K417T at the St. Paul's Hospital Laboratory located within Vancouver Coastal Health Authority[10]; and (4) the commercial Seegene Allplex™ SARS-CoV-2 Variant I Assay (Seegene, Seoul, South Korea) targeting N501Y and E484K at the Kelowna General Hospital Microbiology Laboratory in the Interior Health Authority.

Based on the above, viruses were categorized as follows:

---

[1] Government of Canada. SARS-CoV-2 variants: National definitions, classifications and public health actions. [Accessed 2 June 2021]. Available: https://www.canada.ca/en/public-health/services/diseases/2019-novel-coronavirus-infection/health-professionals/testing-diagnosing-case-reporting/sars-cov-2-variants-national-definitions-classifications-public-health-actions.html
[2] World Health Organization. Tracking SARS-CoV-2 variants. [Accessed 2 June 2021]. Available: https://www.who.int/en/activities/tracking-SARS-CoV-2-variants/
[3] Freed NE, Vlková M, Faisal MB, Silander OK. Rapid and inexpensive whole-genome sequencing of SARS-CoV-2 using 1200 bp tiled amplicons and Oxford Nanopore Rapid Barcoding. Biol Methods Protoc. 2020 Jul 18;5(1):bpaa014.
[4] Galloway SE, Paul P, MacCannell DR, et al. Emergence of SARS-CoV-2 B.1.1.7 Lineage - United States, December 29, 2020-January 12, 2021. MMWR Morb Mortal Wkly Rep. 2021 Jan 22;70(3):95-9.
[5] https://artic.network/ncov-2019
[6] https://github.com/BCCDC-PHL/ncov2019-artic-nf
[7] Rambaut A, Holmes EC, O'Toole A, et al. A dynamic nomenclature proposal for SARS-CoV-2 lineages to assist genomic epidemiology. Nature Microbiology 2020;5:1403-07. Available: https://www.nature.com/articles/s41564-020-0770-5
[8] https://github.com/jts/ncov-tools
[9] Horgan CA, Sbihi H, Jassem A, et al. Optimizing SARS-CoV-2 variant of concern screening: experience from British Columbia, Canada, Early 2021. MedRxiv 2021.03.23.21253520; (Pre-print) doi: https://www.medrxiv.org/content/10.1101/2021.03.23.21253520v1
[10] Matic N, Lowe CF, Ritchie G, et al. Rapid detection of SARS-CoV-2 variants of concern, including B.1.1.28/P.1, British Columbia, Canada. 2021;27(6):1673-1676. doi:10.3201/eid2706.210532.https://wwwnc.cdc.gov/eid/article/27/6/21-0532_article

**Non-VOC**
WGS = non-VOC or
Screening RT-PCR = Negative for N501Y (presumptive)

**B.1.1.7**
WGS = B.1.1.7 or
Screening RT-PCR = Positive for N501Y and positive for 69/70 deletion or positive for N501Y and negative for E484K (presumptive)

**B.1.351***
WGS = B.1.351 or
Screening RT-PCR = Positive for N501Y and negative for K417T (presumptive)

**P.1***
WGS = P.1 or
Screening RT-PCR = Positive for N501Y and positive for K417T (presumptive)

**B.1.617.1/2**
WGS = B.1.617.1/2
Screening RT-PCR = Not established

*Both B.1.351 and P.1 bear the E484K substitution and cannot be distinguished on that basis by RT-PCR screening.

The above algorithm resulted in VOC classification of **1,131/1,226 (92%)** case viruses[1] as shown in the Table S1 below and showing comparable distribution by participant subgroup in Figure S1 below:

**Table S1:** Tally of classified VOC and non-VOC case viruses, by genetic sub-grouping

| | Non-VOC | B.1.1.7 | P.1 and B.1.351 | | | | B.1.617.1/2[1] | Total |
|---|---|---|---|---|---|---|---|---|
| | | | P.1 | 501Y + 417T + | 501Y + and 484K +, 417 SNP not available | B.1.351[1] | | |
| WGS | 65 | 142 | 280 | - | - | 4 | 16 | 510 |
| Screening RT-PCR | 211 | 367 | - | 34 | 12 | - | - | 626 |
| **Total** | 276 | 509 | 326 | | | 4 | 16 | **1,131** |

[1] Excluded from variant-specific VE analyses owing to small sample size

**Variant-specific VE for P.1 was assessed in three ways:**

(1) Confirmed and presumptive P.1 *all* (n=326): defined by WGS (n=280) and screening assay positivity for 501Y and 417T (n=34), also including specimens that were 501Y+484K positive but for which the 417 SNP was unavailable (n=12), the latter presumptively assigned P.1 given a paucity of B.1.351 otherwise detected in BC;

(2) Confirmed and presumptive P.1 *subset* (n=314), defined as above but excluding specimens for which the 417 SNP was not available [NOTE: this group forms the basis of our primary P.1-specific analysis];

(3) Confirmed P.1 (n=280), defined by WGS only.

**VE estimates were similar for all three definitions of P.1** (see **Supplement Table S7** and **Supplement Table S8**). We therefore present definition 2 (n=314 cases) as the primary P.1-specific VE analysis.

[1] Of the 95 case viruses excluded from VOC classification, 6 were subjected to WGS and 89 were not. Of the 6 excluded viruses that were subjected to WGS, 5 were non-VOC that were 484K positive on screening RT-PCR (including 3 designated B.1.525), and one was a presumptive B.1.1.7 on screening RT-PCR not reinforced by WGS (B.1.1). Of the 89 viruses that were not subjected to WGS, screening RT-PCR showed that 2 were 501Y negative (presumptive non-VOC) but 484K positive; 4 were 501Y positive but results for the 417 and 484 SNP were not available; and 85 lacked information on the 501 SNP required for presumptive VOC classification.

**Figure S1:** Distribution of variants of concern (VOC) as identified among genetically characterized viruses contributing to vaccine effectiveness (VE) analyses, adults ≥70 years old, British Columbia (BC), Canada, epidemiological weeks 14-17

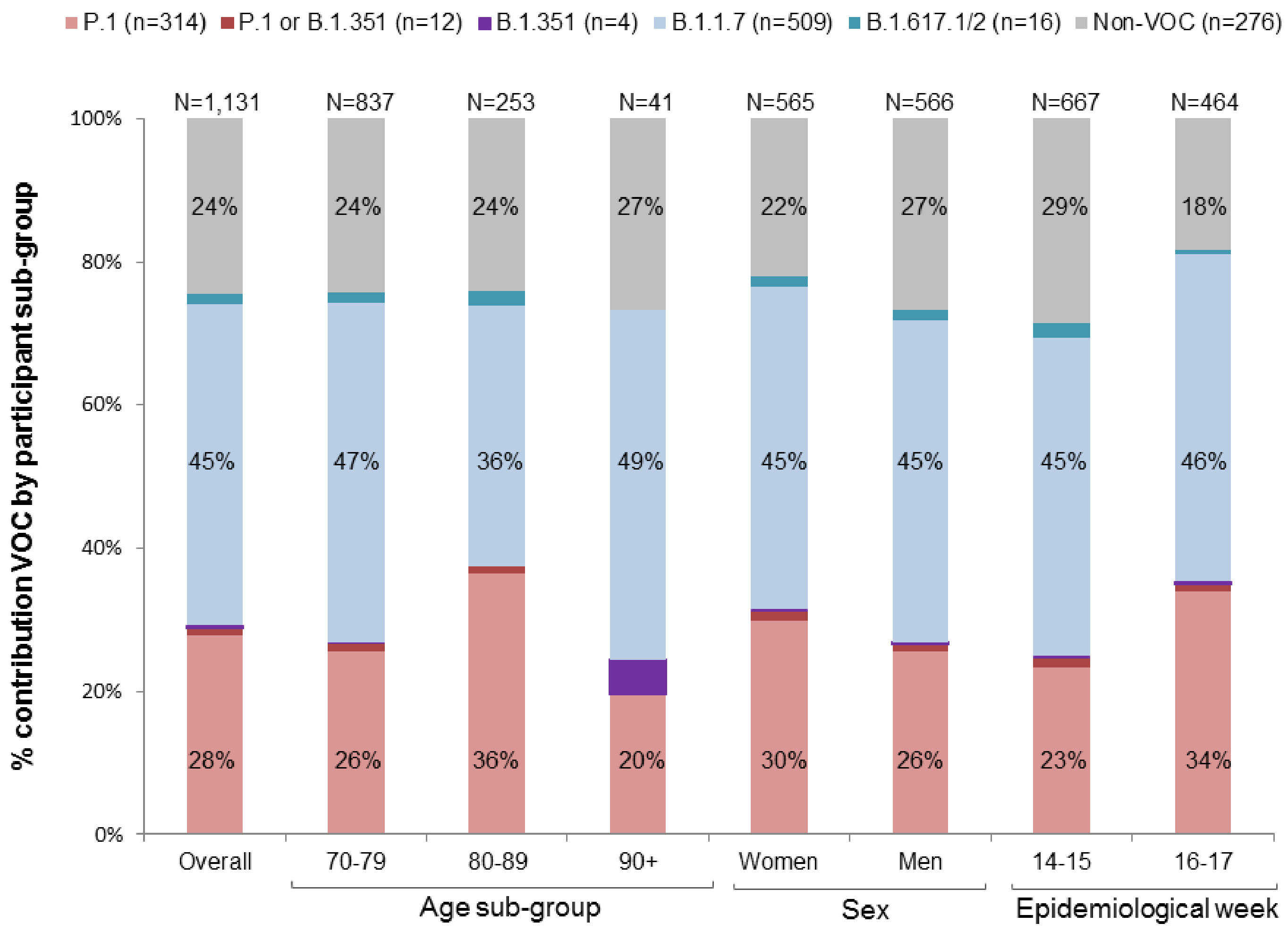

**Supplementary Figure S2**. Specimens from community-dwelling adults ≥70 years old included in vaccine effectiveness (VE) analyses, April 4 (week 14) to May 1 (week 17), 2021, British Columbia (BC), Canada

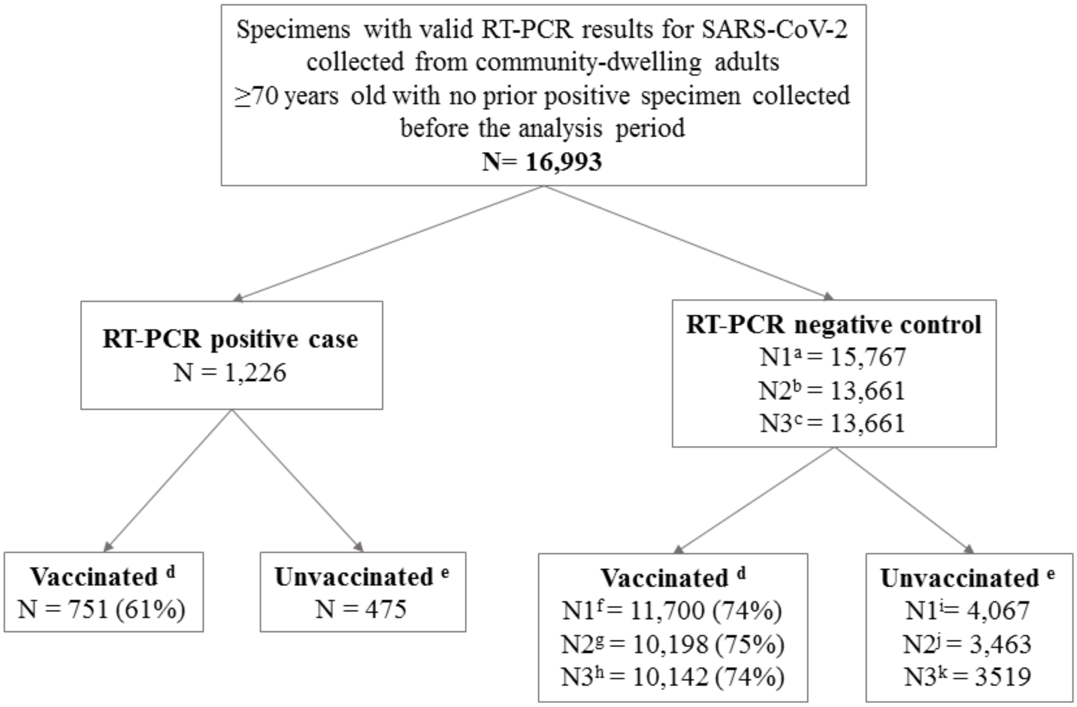

a. Approach 1 for control selection: all negative specimens collected from individuals before the end of the analysis period or becoming a case, whichever occurs first
b. Approach 2 for control selection: one negative specimen (the latest) collected per individual before the end of the analysis period or becoming a case, whichever occurs first
c. Approach 3 for control selection: one randomly selected specimen collected per individual before the end of the analysis period or becoming a case, whichever occurs first
d. Vaccinated on or before the date of specimen collection
e. Not vaccinated on or before the date of specimen collection
f. Vaccinated controls selected as per approach 1
g. Vaccinated controls selected as per approach 2
h. Vaccinated controls selected as per approach 3
i. Unvaccinated controls selected as per approach 1
j. Unvaccinated controls selected as per approach 2
k. Unvaccinated controls selected as per approach 3

**Supplementary Table S2**. Single-dose mRNA vaccine effectiveness by approach for test-negative control selection, adults ≥70 years, BC, Canada

| Interval (DSV) | Case status | By approach for selecting among repeat test-negative control specimens | | | | | | | | | | | |
|---|---|---|---|---|---|---|---|---|---|---|---|---|---|
| | | Approach 1 (all test-negative controls) | | | | Approach 2 (single latest test-negative control) | | | | Approach 3 (single random test-negative control) | | | |
| | | Vacc N (%) | Unvacc N | Crude VE % (95% CI) | Adjusted[1] VE % (95% CI) | Vacc N (%) | Unvacc N | Crude VE % (95% CI) | Adjusted[1] VE % (95% CI) | Vacc N (%) | Unvacc N | Crude VE % (95% CI) | Adjusted[1] VE % (95% CI) |
| Without excluding test-negative specimens collected within three weeks prior to a test-positive specimen | | | | | | | | | | | | | |
| 0-13 | Case | 345 (42%) | 475 | 4 (-11, 17) | 14 (0, 26) | 345 (42%) | 475 | 6 (-9, 19) | 16 (2, 28) | 345 (42%) | 475 | 7 (-8, 20) | 16 (3, 28) |
| | Control | 3078 (43%) | 4067 | | | 2666 (43%) | 3463 | | | 2740 (44%) | 3519 | | |
| 14-20 | Case | 163 (26%) | 475 | 39 (27, 50) | 43 (30, 53) | 163 (26%) | 475 | 41 (29, 51) | 44 (32, 54) | 163 (26%) | 475 | 40 (28, 51) | 45 (33, 54) |
| | Control | 2301 (36%) | 4067 | | | 2014 (37%) | 3463 | | | 2029 (37%) | 3519 | | |
| 21-27 | Case | 110 (19%) | 475 | 57 (47, 65) | 57 (47, 66) | 110 (19%) | 475 | 58 (48, 66) | 59 (48, 67) | 110 (19%) | 475 | 57 (47, 66) | 59 (49, 68) |
| | Control | 2192 (35%) | 4067 | | | 1912 (36%) | 3463 | | | 1909 (35%) | 3519 | | |
| 28-34 | Case | 61 (11%) | 475 | 71 (62, 78) | 69 (59, 77) | 61 (11%) | 475 | 71 (62, 78) | 70 (59, 77) | 61 (11%) | 475 | 70 (61, 78) | 70 (59, 77) |
| | Control | 1790 (31%) | 4067 | | | 1557 (31%) | 3463 | | | 1529 (30%) | 3519 | | |
| 35-41 | Case | 30 (6%) | 475 | 78 (68, 85) | 75 (63, 83) | 30 (6%) | 475 | 78 (69, 85) | 75 (63, 83) | 30 (6%) | 475 | 77 (67, 84) | 75 (63, 83) |
| | Control | 1180 (22%) | 4067 | | | 1016 (23%) | 3463 | | | 983 (22%) | 3519 | | |
| ≥42 | Case | 42 (8%) | 475 | 69 (57, 77) | 63 (48, 74) | 42 (8%) | 475 | 70 (59, 78) | 64 (48, 75) | 42 (8%) | 475 | 67 (54, 76) | 63 (47, 74) |
| | Control | 1150 (22%) | 4067 | | | 1024 (23%) | 3463 | | | 945 (21%) | 3519 | | |
| ≥21 | Case | 243 (34%) | 475 | 67 (61, 72) | 65 (58, 71) | 243 (34%) | 475 | 68 (62, 73) | 66 (59, 72) | 243 (34%) | 475 | 66 (61, 71) | 66 (59, 72) |
| | Control | 6321 (61%) | 4067 | | | 5518 (61%) | 3463 | | | 5373 (60%) | 3519 | | |
| Excluding negative specimens collected within three weeks prior to a test-positive specimen | | | | | | | | | | | | | |
| 0-13 | Case | 345 (42%) | 475 | 4 (-11, 17) | 13 (-1, 26) | 345 (42%) | 475 | 6 (-10, 18) | 16 (2, 28) | 345 (42%) | 475 | 6 (-9, 19) | 16 (2, 28) |
| | Control | 3050 (43%) | 4033 | | | 2640 (43%) | 3434 | | | 2694 (44%) | 3499 | | |
| 14-20 | Case | 163 (26%) | 475 | 40 (27, 50) | 43 (31, 53) | 163 (26%) | 475 | 41 (29, 51) | 44 (32, 54) | 163 (26%) | 475 | 41 (28, 51) | 45 (33, 55) |
| | Control | 2289 (36%) | 4033 | | | 2002 (37%) | 3434 | | | 2018 (37%) | 3499 | | |
| 21-27 | Case | 110 (19%) | 475 | 57 (47, 66) | 58 (47, 66) | 110 (19%) | 475 | 58 (48, 66) | 59 (48, 67) | 110 (19%) | 475 | 58 (47, 66) | 59 (49, 68) |
| | Control | 2186 (35%) | 4033 | | | 1906 (36%) | 3434 | | | 1913 (35%) | 3499 | | |
| 28-34 | Case | 61 (11%) | 475 | 71 (62, 78) | 69 (59, 77) | 61 (11%) | 475 | 72 (63, 78) | 70 (60, 78) | 61 (11%) | 475 | 70 (61, 78) | 70 (60, 78) |
| | Control | 1785 (31% | 4033 | | | 1552 (31%) | 3434 | | | 1520 (30%) | 3499 | | |
| 35-41 | Case | 30 (6%) | 475 | 78 (69, 85) | 75 (63, 83) | 30 (6%) | 475 | 79 (69, 85) | 76 (63, 84) | 30 (6%) | 475 | 77 (67, 84) | 75 (63, 83) |
| | Control | 1177 (23%) | 4033 | | | 1013 (23%) | 3434 | | | 972 (22%) | 3499 | | |
| ≥42 | Case | 42 (8%) | 475 | 69 (57, 77) | 63 (48, 74) | 42 (8%) | 475 | 70 (59, 78) | 64 (48, 75) | 42 (8%) | 475 | 67 (55, 76) | 63 (47, 74) |
| | Control | 1141 (22%) | 4033 | | | 1017 (23%) | 3434 | | | 950 (21%) | 3499 | | |
| ≥21 | Case | 243 (34%) | 475 | 67 (62, 72) | 66 (59, 71) | 243 (34%) | 475 | 68 (62, 73) | 66 (59, 72) | 243 (34%) | 475 | 67 (61, 72) | 66 (59, 72) |
| | Control | 6298 (61%) | 4033 | | | 5497 (62%) | 3434 | | | 5362 (61%) | 3499 | | |

DSV = Days since vaccination, the interval in days between the date of receipt of the first mRNA dose and the date of specimen collection
Vacc = vaccinated; Unvacc = Unvaccinated; VE = vaccine effectiveness; 95% CI = 95% confidence interval

[1] VE estimates adjusted for age group (70-79, 80-89, 90+ years); sex (men, women); epidemiological week (14, 15, 16, or 17); and health authority (HA) (Fraser HA, Interior HA, Northern HA, Vancouver Coastal HA, Vancouver Island HA).

**Supplementary Table S3**. Single-dose mRNA vaccine effectiveness by age subgroup, adults ≥70 years, British Columbia, Canada

| Interval (DSV) | Case status | By age subgroup | | | | | | | | | | | |
|---|---|---|---|---|---|---|---|---|---|---|---|---|---|
| | | 70-79 years | | | | 80-89 years | | | | ≥80 years | | | |
| | | Vacc N (%) | Unvacc N | Crude VE % (95% CI) | Adjusted[1] VE % (95% CI) | Vacc N (%) | Unvacc N | Crude VE % (95% CI) | Adjusted[1] VE % (95% CI) | Vacc N (%) | Unvacc N | Crude VE % (95% CI) | Adjusted[2] VE % (95% CI) |
| 0-13 | Case | 296 (44%) | 384 | 7 (-9, 21) | 16 (1, 28) | 45 (36%) | 80 | 11 (-30, 39) | 6 (-41, 38) | 49 (35%) | 91 | 2 (-40, 32) | 5 (-41, 35) |
| | Control | 2492 (45%) | 3003 | | | 523 (39%) | 825 | | | 586 (36%) | 1064 | | |
| 14-20 | Case | 116 (23%) | 384 | 40 (26, 52) | 46 (32, 57) | 43 (35%) | 80 | 36 (6, 56) | 39 (9, 59) | 47 (34%) | 91 | 29 (-2, 51) | 36 (6, 56) |
| | Control | 1522 (34%) | 3003 | | | 692 (46%) | 825 | | | 779 (42%) | 1064 | | |
| 21-27 | Case | 64 (14%) | 384 | 56 (42, 66) | 61 (47, 71) | 39 (33%) | 80 | 54 (32, 69) | 54 (32, 70) | 46 (34%) | 91 | 49 (27, 65) | 50 (27, 66) |
| | Control | 1127 (27%) | 3003 | | | 880 (52%) | 825 | | | 1065 (50%) | 1064 | | |
| 28-34 | Case | 23 (6%) | 384 | 75 (62, 84) | 77 (64, 85) | 31 (28%) | 80 | 62 (42, 75) | 64 (44, 770 | 38 (29%) | 91 | 58 (38, 71) | 59 (39, 73) |
| | Control | 734 (20% | 3003 | | | 838 (50%) | 825 | | | 1056 (50%) | 1064 | | |
| 35-41 | Case | 13 (3%) | 384 | 70 (47, 83) | 73 (51, 85) | 15 (16%) | 80 | 75 (57, 86) | 75 (55, 86) | 17 (16%) | 91 | 77 (60, 86) | 76 (59, 86) |
| | Control | 335 (10%) | 3003 | | | 626 (43%) | 825 | | | 845 (44%) | 1064 | | |
| ≥42 | Case | 17 (4%) | 384 | 60 (33, 75) | 63 (38, 78) | 18 (18%) | 80 | 65 (40, 79) | 66 (39, 81) | 25 (22%) | 91 | 64 (44, 77) | 64 (39, 78) |
| | Control | 329 (10%) | 3003 | | | 526 (39%) | 825 | | | 821 (44%) | 1064 | | |
| ≥21 | Case | 117 (23%) | 384 | 64 (55, 71) | 67 (59, 74) | 103 (56%) | 80 | 63 (50, 73) | 65 (52, 75) | 126 (58%) | 91 | 61 (49, 71) | 62 (49, 72) |
| | Control | 2530 (46%) | 3003 | | | 2873 (78%) | 825 | | | 3791 (78%) | 1064 | | |

DSV = Days since vaccination, the interval in days between the date of receipt of the first mRNA dose and the date of specimen collection
Vacc = vaccinated; Unvacc = Unvaccinated; VE = vaccine effectiveness; 95% CI = 95% confidence interval

[1] VE estimates adjusted for sex (men, women); epidemiological week (14, 15, 16, or 17); and health authority (HA) (Fraser HA, Interior HA, Northern HA, Vancouver Coastal HA, Vancouver Island HA).

[2] VE estimates adjusted for age group (80-89, 90+ years); sex (men, women); epidemiological week (14, 15, 16, or 17); and health authority (HA) (Fraser HA, Interior HA, Northern HA, Vancouver Coastal HA, Vancouver Island HA).

**Supplementary Table S4**. Single-dose mRNA vaccine effectiveness by sex, adults ≥70 years, British Columbia, Canada

| Interval (DSV) | Case status | By sex | | | | | | | |
|---|---|---|---|---|---|---|---|---|---|
| | | Women | | | | Men | | | |
| | | Vaccinated N (%) | Unvaccinated N | Crude VE % (95% CI) | Adjusted[1] VE % (95% CI) | Vaccinated N (%) | Unvaccinated N | Crude VE % (95% CI) | Adjusted[1] VE % (95% CI) |
| 0-13 | Case | 163 (40%) | 247 | 8 (-13, 25) | 16 (-4, 32) | 182 (44%) | 228 | 0 (-23, 18) | 10 (-11, 28) |
| | Control | 1475 (42%) | 2054 | | | 1603 (44%) | 2013 | | |
| 14-20 | Case | 83 (25%) | 247 | 41 (24, 55) | 44 (28, 57) | 80 (26%) | 228 | 37 (18, 52) | 40 (20, 54) |
| | Control | 1175 (36%) | 2054 | | | 1126 (36%) | 2013 | | |
| 21-27 | Case | 48 (16) | 247 | 65 (51, 74) | 66 (52, 75) | 62 (21%) | 228 | 48 (31, 61) | 47 (28, 62) |
| | Control | 1129 (35%) | 2054 | | | 1063 (35%) | 2013 | | |
| 28-34 | Case | 37 (13%) | 247 | 68 (54, 77) | 67 (52, 78) | 24 (10%) | 228 | 74 (61, 83) | 72 (55, 82) |
| | Control | 960 (32%) | 2054 | | | 830 (29% | 2013 | | |
| 35-41 | Case | 16 (6%) | 247 | 79 (65, 87) | 77 (60, 87) | 14 (6%) | 228 | 78 (61, 87) | 72 (50, 85) |
| | Control | 627 (23%) | 2054 | | | 553 (22%) | 2013 | | |
| ≥42 | Case | 18 (7%) | 247 | 76 (61, 85) | 72 (52, 83) | 24 (10%) | 228 | 60 (39, 74) | 53 (23, 71) |
| | Control | 619 (23%) | 2054 | | | 531 (21%) | 2013 | | |
| ≥21 | Case | 119 (33%) | 247 | 70 (63, 76) | 70 (61, 76) | 124 (35%) | 228 | 63 (54, 71) | 60 (48, 70) |
| | Control | 3341 (62%) | 2054 | | | 2980 (60%) | 2013 | | |

DSV = Days since vaccination, the interval in days between the date of receipt of the first mRNA dose and the date of specimen collection
VE = vaccine effectiveness; 95% CI = 95% confidence interval

[1] VE estimates adjusted for: age group (70-79, 80-89, 90+ years); epidemiological week (14, 15, 16, or 17); and health authority (HA) (Fraser HA, Interior HA, Northern HA, Vancouver Coastal HA, Vancouver Island HA).

**Supplementary Table S5**. Single-dose mRNA vaccine effectiveness by epidemiological week, adults ≥70 years, British Columbia, Canada

| Interval (DSV) | Case status | By epidemiological week (bi-weekly) | | | | | | | |
|---|---|---|---|---|---|---|---|---|---|
| | | Weeks 14-15 | | | | Weeks 16-17 | | | |
| | | Vaccinated N (%) | Unvaccinated N | Crude VE % (95% CI) | Adjusted[1] VE % (95% CI) | Vaccinated N (%) | Unvaccinated N | Crude VE % (95% CI) | Adjusted[2] VE % (95% CI) |
| 0-13 | Case | 228 (41% | 327 | 9 (-9, 24) | 18 (2, 32) | 117 (44%) | 148 | -8 (-39, 17) | 2 (-28, 26) |
| | Control | 2101 (43%) | 2738 | | | 977 (42%) | 1329 | | |
| 14-20 | Case | 85 (21%) | 327 | 41 (25, 54) | 41 (23, 55) | 78 (35%) | 148 | 35 (14, 51) | 45 (25, 59) |
| | Control | 1216 (31%) | 2738 | | | 1085 (45%) | 1329 | | |
| 21-27 | Case | 42 (11%) | 327 | 63 (48, 73) | 59 (42, 71) | 68 (31%) | 148 | 51 (34, 64) | 56 (41, 68) |
| | Control | 945 (26%) | 2738 | | | 1247 (48%) | 1329 | | |
| 28-34 | Case | 24 (7%) | 327 | 66 (47, 77) | 60 (37, 75) | 37 (20% | 148 | 72 (60, 81) | 75 (63, 83) |
| | Control | 583 (18%) | 2738 | | | 1207 (48%) | 1329 | | |
| 35-41 | Case | 6 (2%) | 327 | 84 (63, 93) | 81 (56, 92) | 24 (14%) | 148 | 75 (62, 84) | 73 (57, 83) |
| | Control | 305 (10%) | 2738 | | | 875 (40%) | 1329 | | |
| ≥42 | Case | 5 (2%) | 327 | 74 (37, 90) | 72 (29, 89) | 37 (20%) | 148 | 66 (51, 77) | 63 (45, 75) |
| | Control | 163 (6%) | 2738 | | | 987 (43%) | 1329 | | |
| ≥21 | Case | 77 (19%) | 327 | 68 (58, 75) | 64 (52, 73) | 166 (53%) | 148 | 66 (57, 73) | 67 (58, 74) |
| | Control | 1997 (42%) | 2738 | | | 4324 (76%) | 1329 | | |

DSV = Days since vaccination, the interval in days between the date of receipt of the first mRNA dose and the date of specimen collection
VE = vaccine effectiveness; 95% CI = 95% confidence interval

[1] VE estimates adjusted for age group (70-79, 80-89, 90+ years); sex (men, women); epidemiological week (14, 15); and health authority (HA) (Fraser HA, Interior HA, Northern HA, Vancouver Coastal HA, Vancouver Island HA).

[2] VE estimates adjusted for age group (70-79, 80-89, 90+ years); sex (men, women); epidemiological week (16,17); and health authority (HA) (Fraser HA, Interior HA, Northern HA, Vancouver Coastal HA, Vancouver Island HA).

**Supplementary Table S6**. Single-dose mRNA vaccine effectiveness by mRNA product, adults ≥70 years, British Columbia, Canada

| Interval (DSV) | Case status | By mRNA product (Pfizer-BioNTech; Moderna) | | | | | | | |
|---|---|---|---|---|---|---|---|---|---|
| | | Pfizer-BioNTech | | | | Moderna | | | |
| | | Vaccinated N (%) | Unvaccinated N | Crude VE % (95% CI) | Adjusted[1] VE % (95% CI) | Vaccinated N (%) | Unvaccinated N | Crude VE % (95% CI) | Adjusted VE % (95% CI) |
| 0-13 | Case | 288 (38) | 475 (62) | 4 (-12, 18) | 14 (-1, 26) | 57 (11) | 475 (89) | 3 (-30, 27) | 11 (-20, 34) |
| | Control | 2575 (39) | 4067 (61) | | | 503 (11) | 4067 (89) | | |
| 14-20 | Case | 143 (23) | 475 (77) | 37 (23, 48) | 42 (28, 52) | 20 (4) | 475 (96) | 53 (25, 70) | 49 (19, 68) |
| | Control | 1940 (32) | 4067 (68) | | | 361 (8) | 4067 (92) | | |
| 21-27 | Case | 102 (18) | 475 (82) | 53 (42, 62) | 54 (41, 63) | 8 (2) | 475 (98) | 79 (58, 90) | 78 (55, 89) |
| | Control | 1864 (31) | 4067 (69) | | | 328 (7) | 4067 (93) | | |
| 28-34 | Case | 53 (10) | 475 (90) | 71 (61, 78) | 69 (58, 77) | 8 (2) | 475 (98) | 71 (41, 86) | 68 (34, 85) |
| | Control | 1555 (28) | 4067 (72) | | | 235 (5) | 4067 (95) | | |
| 35-41 | Case | 25 (5) | 475 (95) | 78 (67, 86) | 75 (61, 84) | 5 (1) | 475 (99) | 78 (47, 91) | 75 (39, 90) |
| | Control | 983 (19) | 4067 (81) | | | 197 (5) | 4067 (95) | | |
| ≥42 | Case | 35 (7) | 475 (93) | 70 (58, 79) | 66 (51, 77) | 7 (1) | 475 (99) | 60 (14, 81) | 42 (-28, 74) |
| | Control | 1001 (20) | 4067 (80) | | | 149 (4) | 4067 (96) | | |
| ≥21 | Case | 215 (31) | 475 (69) | 66 (60, 71) | 64 (57, 71) | 28 (6) | 475 (94) | 74 (61, 82) | 71 (56, 81) |
| | Control | 5408 (57) | 4067 (43) | | | 913 (18) | 4067 (82) | | |

[1] VE estimates adjusted for age group (70-79, 80-89, 90+ years); sex (men, women); epidemiological week (14, 15, 16, or 17); and health authority (HA) (Fraser HA, Interior HA, Northern HA, Vancouver Coastal HA, Vancouver Island HA).

**Supplementary Table S7**. Single-dose mRNA vaccine effectiveness by variant of concern (VOC) status: non-VOC, B.1.1.7 and P.1, adults ≥70 years, British Columbia, Canada

| Interval (DSV) | Case Status | Non-VOC[1] (n=276 cases) | | | | B.1.1.7[1] (n=509 cases) | | | | P.1[1,2] (n=314 cases) | | | |
|---|---|---|---|---|---|---|---|---|---|---|---|---|---|
| | | Vaccinated N (%) | Unvaccinated N | Crude VE % (95% CI) | Adjusted[3] VE % (95% CI) | Vaccinated N (%) | Unvaccinated N | Crude VE % (95% CI) | Adjusted[2] VE % (95% CI) | Vaccinated N (%) | Unvaccinated N | Crude VE % (95% CI) | Adjusted[2] VE % (95% CI) |
| 0-13 | Case | 84 (40%) | 126 | 12 | 17 | 140 (41%) | 200 | 8 | 18 | 81 (43%) | 109 | 2 | 11 |
| | Control | 3078 (43%) | 4067 | (-17, 33) | (-10, 38) | 3078 (43%) | 4067 | (-15, 26) | (-3, 34) | 3078 (43%) | 4067 | (-31, 27) | (-19, 34) |
| 14-20 | Case | 24 (16%) | 126 | 66 | 63 | 66 (25%) | 200 | 42 | 48 | 53 (33%) | 109 | 14 | 24 |
| | Control | 2301 (36%) | 4067 | (48, 78) | (42, 77) | 2301 (36%) | 4067 | (23, 56) | (30, 61) | 2301 (36%) | 4067 | (-20, 38) | (-7, 46) |
| 21-27 | Case | 15 (11%) | 126 | 78 | 74 | 52 (21%) | 200 | 52 | 55 | 30 (22%) | 109 | 49 | 52 |
| | Control | 2192 (35%) | 4067 | (62, 87) | (55, 85) | 2192 (35%) | 4067 | (34, 65) | (37, 67) | 2192 (35%) | 4067 | (23, 66) | (26, 69) |
| 28-34 | Case | 12 (9%) | 126 | 78 | 70 | 22 (10%) | 200 | 75 | 76 | 18 (14%) | 109 | 62 | 66 |
| | Control | 1790 (31%) | 4067 | (61, 88) | (43, 84) | 1790 (31%) | 4067 | (61, 84) | (62, 85) | 1790 (31%) | 4067 | (38, 77) | (42, 80) |
| 35-41 | Case | 9 (7%) | 126 | 75 | 65 | 11 (5%) | 200 | 81 | 81 | 9 (8%) | 109 | 72 | 68 |
| | Control | 1180 (22%) | 4067 | (51, 88) | (27, 83) | 1180 (22%) | 4067 | (65, 90) | 64, 90) | 1180 (22%) | 4067 | (44, 86) | (35, 85) |
| ≥42 | Case | 6 (5%) | 126 | 83 | 72 | 18 (8%) | 200 | 68 | 67 | 14 (11%) | 109 | 55 | 53 |
| | Control | 1150 (22%) | 4067 | (62, 93) | (33, 88) | 1150 (22%) | 4067 | (48, 80) | (44, 81) | 1150 (22%) | 4067 | (20, 74) | (12, 75) |
| ≥21 | Case | 42 (25%) | 126 | 79 | 72 | 103 (34%) | 200 | 67 | 67 | 71 (39%) | 109 | 58 | 61 |
| | Control | 6321 (61%) | 4067 | (70, 85) | (58, 81) | 6321 (61%) | 4067 | (58, 74) | (57, 75) | 6321 (61%) | 4067 | (43, 69) | (45, 72) |

DSV = Days since vaccination, the interval in days between the date of receipt of the first mRNA dose and the date of specimen collection
VOC = variant of concern; VE = vaccine effectiveness; 95% CI = 95% confidence interval

[1] Defined as in Supplementary Material S1.

[2] Confirmed and presumptive P.1 subset (n=314) defined by whole genome sequencing (WGS) (n=280) and screening real-time reverse transcriptase polymerase chain reaction (RT-PCR) assay positivity for 501Y and 417T (n=34), excluding specimens that were 501Y positive and 484K positive but for which the 417 SNP was not available (n=12). See Supplementary Material S1.

[3] VE estimates adjusted for age group (70-79, 80-89, 90+ years); sex (men, women); epidemiological week (14, 15, 16, or 17); and health authority (HA) (Fraser HA, Interior HA, Northern HA, Vancouver Coastal HA, Vancouver Island HA).

**Supplementary Table S8**. Single-dose mRNA vaccine effectiveness: P.1 sensitivity analysis, adults ≥70 years, British Columbia, Canada

| Interval (DSV) | Case Status | Confirmed and presumptive P.1 (n=326 P.1 cases)[1] | | | | Confirmed P.1 only (n=280 P.1 cases)[2] | | | |
|---|---|---|---|---|---|---|---|---|---|
| | | Vacc N (%) | Unvacc N | Crude VE % (95% CI) | Adjusted[3] VE % (95% CI) | Vacc N (%) | Unvacc N | Crude VE % (95% CI) | Adjusted[4] VE % (95% CI) |
| 0-13 | Case | 85 (43%) | 112 | 0 (-33, 25) | 9 (-22, 32) | 72 (42%) | 98 | 3 (-32, 29) | 10 (-23, 34) |
| | Control | 3078 (43%) | 4067 | | | 3078 (43%) | 4067 | | |
| 14-20 | Case | 56 (33%) | 112 | 12 (-22, 36) | 22 (-9, 44) | 47 (32%) | 98 | 15 (-20, 40) | 25 (-7, 48) |
| | Control | 2301 (36%) | 4067 | | | 2301 (36%) | 4067 | | |
| 21-27 | Case | 31 (22%) | 112 | 49 (23, 66) | 51 (25, 68) | 24 (20%) | 98 | 55 (29, 71) | 59 (35, 75) |
| | Control | 2192 (35%) | 4067 | | | 2192 (35%) | 4067 | | |
| 28-34 | Case | 19 (15%) | 112 | 61 (37, 76) | 65 (40, 79) | 18 (16%) | 98 | 58 (31, 75) | 64 (39, 79) |
| | Control | 1790 (31%) | 4067 | | | 1790 (31%) | 4067 | | |
| 35-41 | Case | 9 (7%) | 112 | 72 (45, 86) | 69 (35, 85) | 8 (8%) | 98 | 72 (42, 86) | 71 (37, 87) |
| | Control | 1180 (22%) | 4067 | | | 1180 (22%) | 4067 | | |
| ≥42 | Case | 14 (11%) | 112 | 56 (23, 75) | 53 (13, 75) | 13 (12%) | 98 | 53 (16, 74) | 54 (13, 76) |
| | Control | 1150 (22%) | 4067 | | | 1150 (22%) | 4067 | | |
| ≥21 | Case | 73 (40%) | 112 | 58 (44, 69) | 60 (44, 72) | 63 (39%) | 98 | 59 (43, 70) | 63 (47, 74) |
| | Control | 6321 (61%) | 4067 | | | 6321 (61%) | 4067 | | |

DSV = Days since vaccination, the interval in days between the date of receipt of the first mRNA dose and the date of specimen collection
Vacc = vaccinated; Unvacc = Unvaccinated; VE = vaccine effectiveness; 95% CI = 95% confidence interval

[1] Confirmed and all presumptive P.1 (n=326) defined by whole genome sequencing (WGS) (n=280) and screening real-time reverse transcription polymerase chain reaction (RT-PCR) assay positivity for 501Y and 417T (n=34), and also including specimens that were 501Y positive and 484K positive but for which the 417 SNP was not available (n=12) on the added assumption they are most likely P.1 (rather than B.1.351). See **Supplementary Material S1**.

[2] Confirmed P.1 (n=280), defined by WGS only. See **Supplementary Material S1**.

[3] VE estimates adjusted for age group (70-79, 80-89, 90+ years); sex (men, women); epidemiological week (14, 15, 16, or 17; and health authority (HA) (Fraser HA, Interior HA, Northern HA, Vancouver Coastal HA, Vancouver Island HA).
